# Perturbation-based balance training using trips and slips can reduce fall injuries in older adults: The SafeTrip randomised controlled trial

**DOI:** 10.64898/2026.02.20.26346756

**Authors:** Yoshiro Okubo, Steven Phu, Carly Chaplin, Cameron Hicks, Eve Coleman, Peter Humburg, Paula Santiago Martinez, Stephen R Lord

**Author notes:** Corresponding Author: Yoshiro Okubo, PhD, Neuroscience Research Australia, 139 Barker Street, Randwick, New South Wales, Australia 2031. Dual first authors (Yoshiro Okubo, Steven Phu). Portions of this work were presented at the International Society for Gait and Posture Research (ISPGR) 2025 Conference, Maastricht, the Netherlands. **Data Availability**: Access to the datasets analysed for this study is subjected to approval by the University of New South Wales Human Research Ethics Committee. Requests to access the datasets should be directed to.

## Abstract

**BACKGROUND:** Fall injuries in older adults are devastating and often caused by impaired reactive balance to unexpected trips and slips, which conventional exercise programs do not target. This study examined whether a low-dose perturbation balance training (PBT) program among older adults can improve balance recovery following trips and slips and reduce falls and fall injuries.

**METHODS:** 111 older adults (65+ years) were randomised into an intervention or control group. The intervention group undertook one weekly PBT session for three weeks on the Trip and Slip Walkway, followed by three-monthly PBT booster sessions over one year, for a total of six sessions. The control group received an educational booklet. Blinded staff assessed laboratory-falls induced by a trip and slip with a safety harness at baseline and one year. Number of falls and fall injuries in daily life were collected weekly for one year.

**RESULTS:** Compared to the control group, the intervention group experienced a 26% reduction in laboratory falls at 12 months (RR = 0.74; 95% CI: 0.54, 0.99; *P* = .040) but not different in number of falls, trip-and slip-encounters in daily life. However, fall-related injuries were reduced by 57% (rate ratio = 0.43; 95% CI: 0.19, 0.94, *P* = .024) over one year. A reduction in falls occurred within the first three months, with greater benefit among participants who completed at least three training sessions.

**CONCLUSIONS:** A low-dose PBT program can improve reactive balance over 12 months and reduced injurious falls by 57%, with benefits likely due to enhanced reactive balance rather than proactive gait strategies. Older adults may require at least three sessions to achieve meaningful fall reduction, with periodic booster sessions to sustain benefits. Incorporating PBT into exercise programs may enhance their efficacy in preventing falls and fall injuries in daily life.

**Key Points:** A low-dose perturbation-based training program (six sessions over 12 months) improved reactive balance at 12 months and reduced injurious falls by 57%.

Benefits are likely due to task-specific improvements in reactive balance against trips and slips rather than proactive gait strategies or other risk factors.

Incorporating PBT into exercise programs may improve their efficacy in preventing falls and fall injuries in daily life.

**Why does this paper matter?:** Falls are the leading cause of injury-related hospitalization and loss of independence in older adults. By targeting reactive balance—an ability neglected by conventional exercise programs—it offers a novel, evidence-based approach to enhance fall prevention and reduce injuries.

## INTRODUCTION

Falls and fall-related injuries are a major concern, affecting one in three older adults and imposing significant physical, psychological, and economic burdens on individuals, their families, and society.^1^ Tripping over an object or uneven surface and slipping on a slippery surface while walking are the most common causes of falls,^2^ ^3^ and fall-related hospitalisations.^4^ Although conventional exercise interventions address strength and balance impairments ^5^ ^6^, they do not directly target the capacity to respond to trips and slips - an ability that depends on reactive balance. Increasing evidence suggests that perturbation-based balance training (PBT) can address this important risk factor for falls in older adults.^7^

PBT targets reactive balance by providing repeated exposure to unpredictable postural perturbations ^8^ and has been shown to reduce daily-life falls by approximately 39–48%.^9–11^ Although the most effective approach to PBT is unclear, task-specific perturbations that closely resemble real-life challenges may offer the greatest translation to prevent falls.^12^ Treadmill-based PBT are space efficient and easily administered,^13–16^ however, do not adequately replicate a trip event nor sufficiently de-stabilise individuals to elicit trip recovery responses.^17^ ^18^ Consequently, motor adaptations developed in response to treadmill perturbations do not transfer well to obstacle trips,^19^ ^20^ raising concerns regarding translation to real-life trips. Translating treadmill-based slip perturbations to real-life is also challenging, as highlighted in the findings from two randomised controlled trials (RCTs), where a single training session of low-friction slips reduced falls by 50% over the following year,^21^ but a single training session of treadmill-based training did not.^22^ This highlights the importance of conducting PBT using obstacle-trips and low-friction slips to optimise training improvements in reactive balance in older adults.^23^

The long-term effects of PBT are also unclear. A recent PBT trial in older adults suggested that adaptations to a low-dose (4–6 weeks) PBT training program may be retained for only three months^15^ and re-training, or booster sessions can restore training effects.^24^ Therefore, we aimed to determine whether a low-dose PBT program comprised of an initial three-week training period and three-monthly re-training sessions could improve balance recovery following trips and slips and reduce falls and fall injuries in older adults. We hypothesised whether low-dose PBT comprising repeated slips and trips, followed by 3-monthly re-training sessions would: (1) improve reactive balance at 12 months in older adults; (2) reduce near falls (trips and slips), falls, and fall-related injuries in daily life over 12 months; and (3) reduce concerns about falling, improve physiological and cognitive fall risk factors, and increase fall risk awareness. Such findings would provide insights into the task specificity required for effective PBT and strategies to sustain its efficacy to prevent falls in older adults.

## METHODS

### Design, ethics and registration

This was an assessor- and analyst-blinded, two-arm parallel, superiority RCT conducted at Neuroscience Research Australia, Randwick, Australia from 2020-2024. The trial protocol was approved by the University of New South Wales Human Research Ethics Committee (HC210350) and prospectively registered at ClinicalTrials.gov (NCT04602858).

### Study participants

Participants were recruited through research volunteer databases (NeuRA, Join-Us), *Weekend Notes* and word of mouth. Eligible participants were ≥65 years, community-dwelling, and able to walk 500 □ m unaided. Exclusion criteria were a diagnosed neurological condition, a medical condition that prevented exercise such as severe pain, medical advice against exercise, or a history of lower limb, pelvic, or vertebral fractures, or lower limb joint replacements within the past six months. All participants provided written informed consent.

### Sample size

One hundred and twenty participants (two groups of 60) was estimated to be sufficient to detect a 40% difference in the rate of laboratory falls using Poisson regression ^23^ at 12 months with a 20% dropout rate, alpha error of 0.05 and power of 0.8. This sample size was also sufficient for all other analyses involving continuously scored measures except for daily-life falls (secondary outcome) which may require a greater sample size.

### Randomisation and blinding

Participants were randomly assigned in a 1:1 ratio to either the intervention or control group. A random sequence with random block sizes was prepared by an independent researcher using the NeuRA Blinders software. To maintain allocation concealment, randomisation occurred only after each participant completed their baseline assessment. All follow_up assessments were conducted by blinded assessors who were not involved in delivering the intervention.

### Procedure

Usual gait speed, step length and cadence were assessed three times over a 8-metre course using a stopwatch. Participants fitted with a fall-arrest harness (at least 5cm clearance between the knees and the floor) and protective guards walked on the walkway at 90% of their usual gait speed by stepping on tiles placed at 95% of their usual step length and to a metronome set to 95% of usual cadence. The same gait parameters were used for training sessions and at assessments. At least 10 unperturbed walks were completed prior to exposure to the first perturbation test, and at least one washout walk was included between perturbations, delivered at different, unknown locations. Participants were informed that hazards may occur anywhere on the walkway and instructed to walk consistently in accordance with the stepping tiles and metronome beat^23^. Trips were induced using a hidden 14cm height board that flipped up from the walkway at around mid-swing. Slips were induced using a movable tile on two hidden low-friction rails with linear bearings that slid forward on foot contact for up to 70cm. All participants experienced one trip and one slip at baseline and 12 months.^25^

### Interventions

#### Intervention group

The intervention group undertook a low-dose PBT comprised of three 50-minute sessions over three weeks,^23^ followed by re-training sessions every three months at months 3, 6 and 9 (±3-month due to COVID-19 lockdowns). Each session was supervised by an Exercise Physiologist/s and began with a warm_up routine that included dynamic and static stretching.

PBT sessions aimed to improve balance recovery by delivering at least 15 trips and 15 slips whilst walking on the walkway. Sessions progressed in unpredictability and breaks were provided as required. Trips and slips could be induced at various locations (left or right side and beginning, middle or end of the walkway). Trips could also be induced at various points during swing (20-80%) and the slip distances could be adjusted (10-70cm). Unpredictability was progressed based on each participants’ anxiety, perceived difficulty, and performance. Training progressed from “predictable perturbations” of known type, timing, and/or location (e.g. trips occurring in the middle of the walkway) to “unpredictable perturbations” of unknown type, timing, and/or location (e.g. trips, slips or a washout trial occurring anywhere along the walkway and at any time). Where necessary (e.g. due to anxiety), participants were taught optimal reactive stepping patterns in a staged manner, beginning with immediate foot elevation following foot-obstacle contact and progressing to intentional obstacle strikes and finally, unexpected perturbations, where the obstacle suddenly popped up (Appendix A).

Participant anxiety and perceived difficulty levels at the end of each session were recorded using 10-point scales (1: no anxiety at all/easy, 10: extreme anxiety/too hard) for analysis. Exercise enjoyment was assessed using the Physical Activity Enjoyment Scale (PACES).^26^

#### Control group

Both the control and intervention groups received the “Staying Active and On Your Feet” educational booklet on fall prevention, which covers topics such as exercise, diet, mobility, vision, footwear, medications, and home safety (Appendix B).

### Outcomes

#### Reactive balance: Falls from induced slips and trips

The primary outcome was reactive balance, assessed as fall incidence after a slip and trip (described in procedures section) in the laboratory at the 12-month assessment. A fall occurred when the harness supported load exceeded 30% of the person’s body weight following a slip or a trip.^27^

#### Falls in daily life

Near falls involving trips and slips, falls and fall injuries in daily life were prospectively recorded using a falls diary and weekly reports via SMS surveys (REDCap/Twilio) for 52 weeks from randomisation. A trip was defined as catching one’s foot on something causing a loss of balance. A slip was defined as the sliding of one’s foot on a surface causing a loss of balance. A fall was defined as an unexpected event in which the participant comes to rest on the ground, floor or lower level.^28^ Fall-related injuries including soft tissue injuries, cuts, lacerations and fracture were recorded if they reported a fall within the past week.

#### Functional, neuropsychological and behavioural assessments

Fall risk summary scores were calculated using the Physiological Profile Assessment (PPA) short form.^29^ Volitional stepping was assessed using the Choice Stepping Reaction Time (CSRT) test^30^ ^31^ using a monitor and a custom step mat (150 × 90 cm; 2 stance panels, 6 stepping panels). Participants completed 24 random trials, and mean reaction time (ms) from stimulus to foot contact was recorded. Anxiety was assessed using the 7-item General Anxiety Disorder scale (GAD7).^32^ Concern about falling was assessed using the 16-item Falls Efficacy Scale – International (FES-I).^33^ The Trail Making Test (TMT) scores (Parts B–A) were used to assess executive function.^34^ Fall risk awareness and behaviours were assessed using the 24-item Fall Behavioural Scale (FaB).^35^ Total physical activity (hours per week) in the past 3 months was estimated using the Incidental and Planned Exercise Questionnaire (IPEQ).^36^

#### Baseline descriptors

Baseline participant characteristics included age, gender, height, weight, body mass index (BMI), diagnosis of osteoporosis, anxiety disorder, pain (back, hip, knees and feet), use of a walking stick, fear of falling, falls in the past year, the Mini-Addenbrooke’s Cognitive Examination (Mini-ACE), the Brief Pain Inventory, years of education and the Patient Health Questionnaire (PHQ-9).

### Statistical analysis

Primary analyses were conducted with an intention-to-treat (ITT) approach. Continuous data missing at random were imputed using multiple imputation with 50 iterations.^37^ A complier average causal effect (CACE) analysis, with compliance defined by completing at least the initial three training sessions, was conducted. Normality of residuals were confirmed via Q-Q plots, and homogeneity of variance and linearity assumptions were assessed via residual plots. The continuous measurements at post-intervention were compared between the groups using linear regression while adjusting for baseline values. Modified Poisson regression was employed, with trip- or slip-induced falls in the laboratory as the dependent (binary) variable, group allocation as the independent variable while adjusting for baseline laboratory falls.^38^ Negative binomial regression (NBR) analyses were conducted with daily-life fall events (trip/slip encounters, falls, trip falls, slip falls, and fall injuries) as the dependent (count) variable, group allocation as an independent variable, and baseline fall history as a covariate.^38^ To assess retention effects, the 12-month follow-up period was divided into four quarters (Q1–Q4) for analysing NBRs on total falls. Additionally, anxiety, perceived difficulty and enjoyment over the six training sessions were compared using a linear mixed-effects model. IBM SPSS 27 (IBM Corp., NY, USA) was used and *P* < .05 was considered statistically significant. A statistical analysis plan is found in Appendix C.

## RESULTS

### Participant characteristics

Of the 135 older adults expressing interest in participating in the trial, 24 did not attend the baseline assessment. The remaining 111 participants were randomised into the intervention (n=54) and control (n=57) groups and included in the primary ITT analyses (Figure 1). The average age of participants was 73.2±5.4yrs (range 65–91), with height of 163.9±10.2cm, weight of 72.3±13.5kg, and BMI of 26.9±4.9kg/m². Baseline characteristics are shown in Table 1.

**Figure 1.**
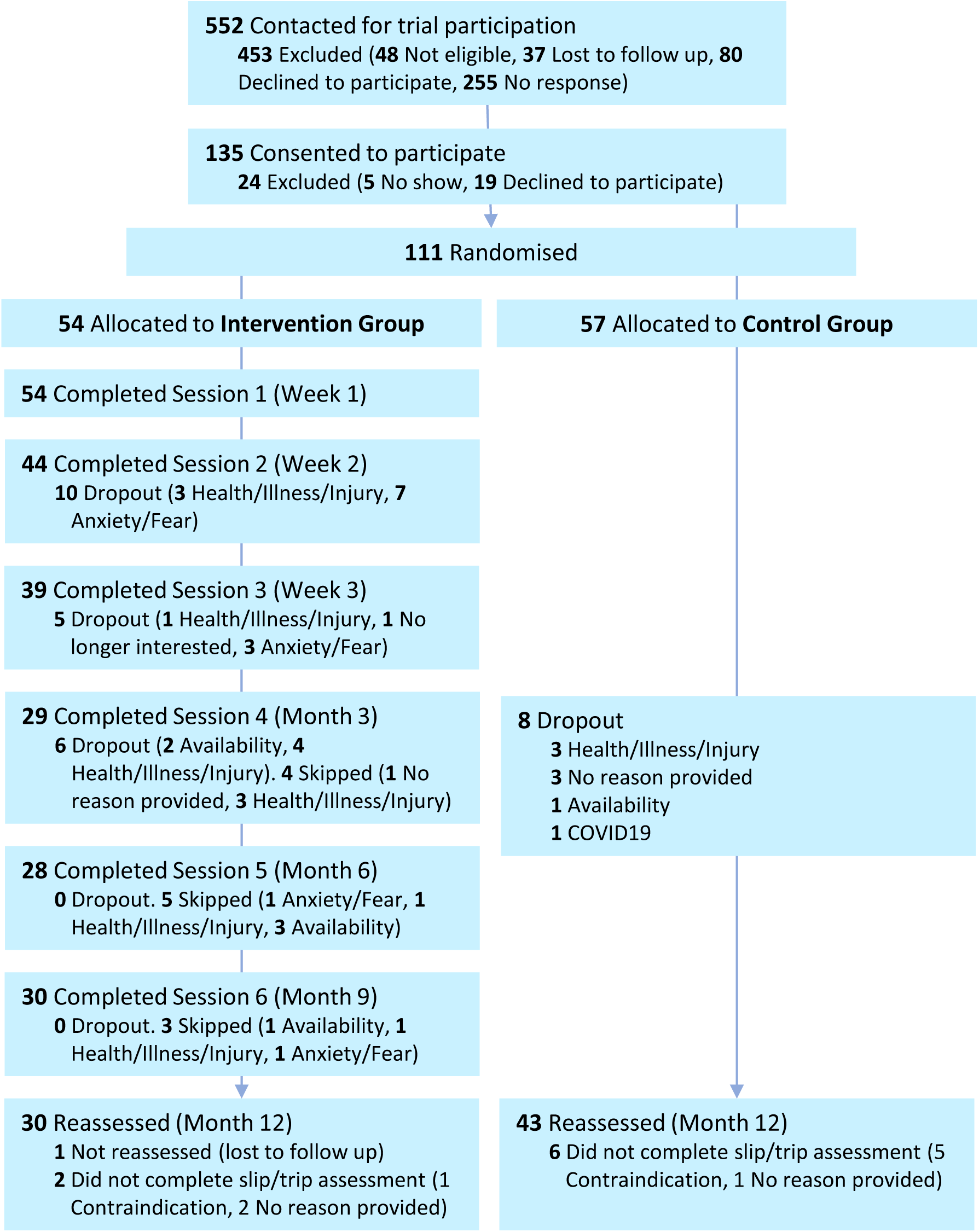
Participant flow in the SafeTrip trial.

**Table 1.**
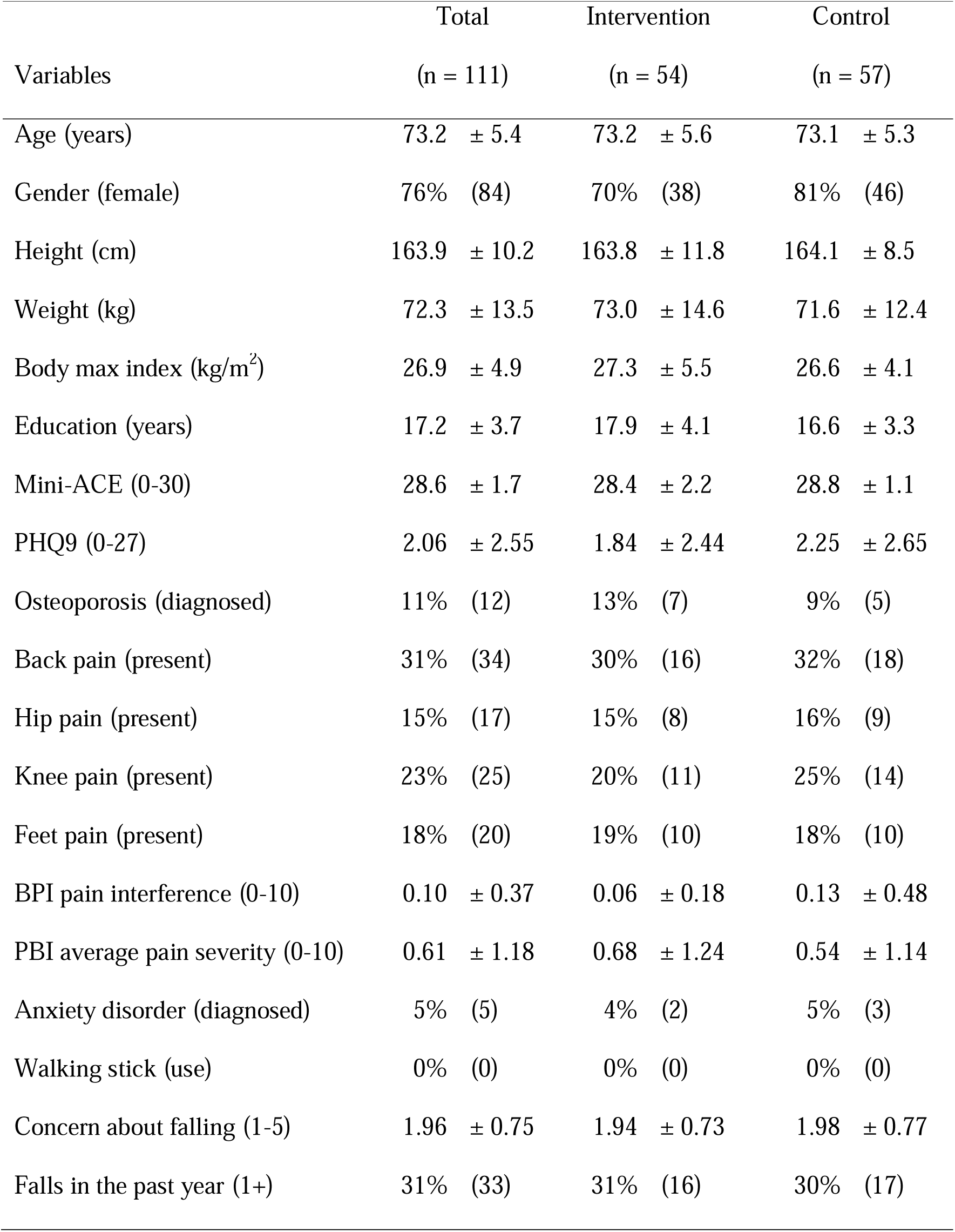
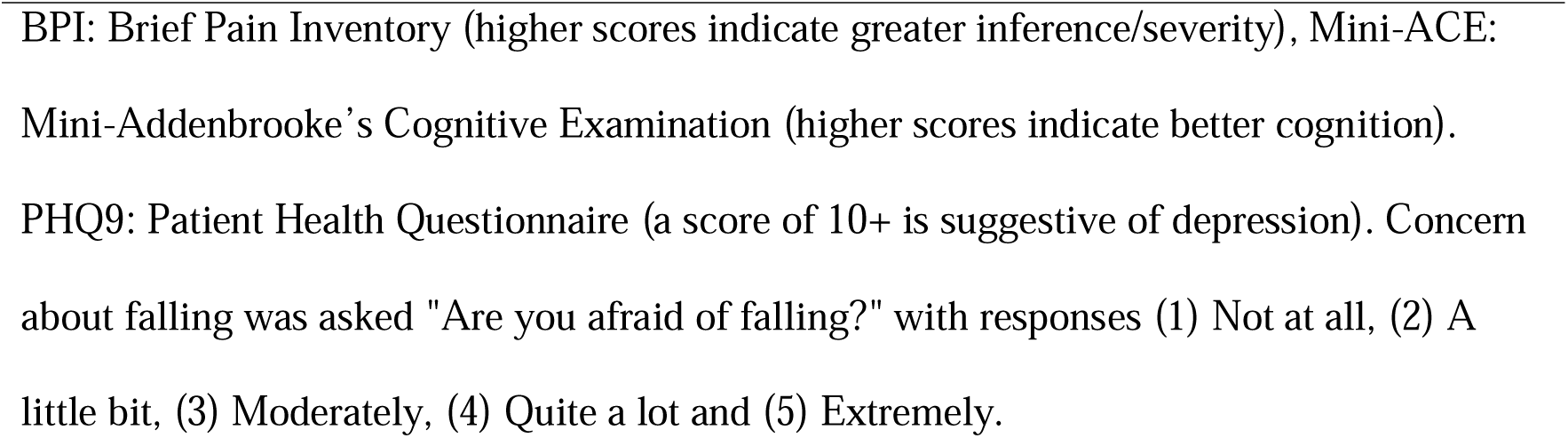
Participant characteristics at baseline.

| Variables | Total<br>(n = 111) | Intervention<br>(n = 54) | Control<br>(n = 57) |
| --- | --- | --- | --- |
| Age (years) | 73.2 ± 5.4 | 73.2 ± 5.6 | 73.1 ± 5.3 |
| Gender (female) | 76% (84) | 70% (38) | 81% (46) |
| Height (cm) | 163.9 ± 10.2 | 163.8 ± 11.8 | 164.1 ± 8.5 |
| Weight (kg) | 72.3 ± 13.5 | 73.0 ± 14.6 | 71.6 ± 12.4 |
| Body mass index (kg/m<sup>2</sup>) | 26.9 ± 4.9 | 27.3 ± 5.5 | 26.6 ± 4.1 |
| Education (years) | 17.2 ± 3.7 | 17.9 ± 4.1 | 16.6 ± 3.3 |
| Mini-ACE (0-30) | 28.6 ± 1.7 | 28.4 ± 2.2 | 28.8 ± 1.1 |
| PHQ9 (0-27) | 2.06 ± 2.55 | 1.84 ± 2.44 | 2.25 ± 2.65 |
| Osteoporosis (diagnosed) | 11% (12) | 13% (7) | 9% (5) |
| Back pain (present) | 31% (34) | 30% (16) | 32% (18) |
| Hip pain (present) | 15% (17) | 15% (8) | 16% (9) |
| Knee pain (present) | 23% (25) | 20% (11) | 25% (14) |
| Feet pain (present) | 18% (20) | 19% (10) | 18% (10) |
| BPI pain interference (0-10) | 0.10 ± 0.37 | 0.06 ± 0.18 | 0.13 ± 0.48 |
| PBI average pain severity (0-10) | 0.61 ± 1.18 | 0.68 ± 1.24 | 0.54 ± 1.14 |
| Anxiety disorder (diagnosed) | 5% (5) | 4% (2) | 5% (3) |
| Walking stick (use) | 0% (0) | 0% (0) | 0% (0) |
| Concern about falling (1-5) | 1.96 ± 0.75 | 1.94 ± 0.73 | 1.98 ± 0.77 |
| Falls in the past year (1+) | 31% (33) | 31% (16) | 30% (17) |

BPI: Brief Pain Inventory (higher scores indicate greater inference/severity), Mini-ACE: Mini-Addenbrooke's Cognitive Examination (higher scores indicate better cognition). PHQ9: Patient Health Questionnaire (a score of 10+ is suggestive of depression). Concern about falling was asked "Are you afraid of falling?" with responses (1) Not at all, (2) A little bit, (3) Moderately, (4) Quite a lot and (5) Extremely.

### Adherence and adverse events

The session attendance rates were 85% during the initial three weeks and 54% during the booster sessions. Over the study period, 22 intervention participants dropped out due to anxiety or fear (n=12), illness or injury (n=8), loss of interest (n=1) and unavailability (n=1). Eight control participants dropped out due to illness or injury (n=3), unavailability (n=1), COVID-19 infection (n=1) and unspecified reasons (n=3). During the first two training sessions, six adverse events were reported: thigh pain (n=1), ankle/knee pain (n=1), groin pain (n=2), a grazed foot (n=1), and a skin tear from electrode removal (n=1). Three participants withdrew due to these events, while the remaining three resumed training once their issues had resolved.

### Anxiety, perceived difficulty and enjoyment

Anxiety significantly changed over the training sessions (F(5, 155.0) = 2.70, *P* = .023) (Figure 2), with the highest levels in session 1 (mean: 4.98, 95% CI: 4.40, 5.56) and the lowest in session 6 (mean: 3.86, 95% CI: 3.12, 4.59). Perceived difficulty did not change (*P* = .216). Enjoyment also significantly changed across sessions (F(5, 185.6) = 7.92, *P* < .001), with the highest in session 1 (mean: 35.7, 95% CI: 33.8, 37.6) and reaching the lowest level in session 6 (mean: 29.0, 95% CI: 26.5, 31.5).

**Figure 2.**
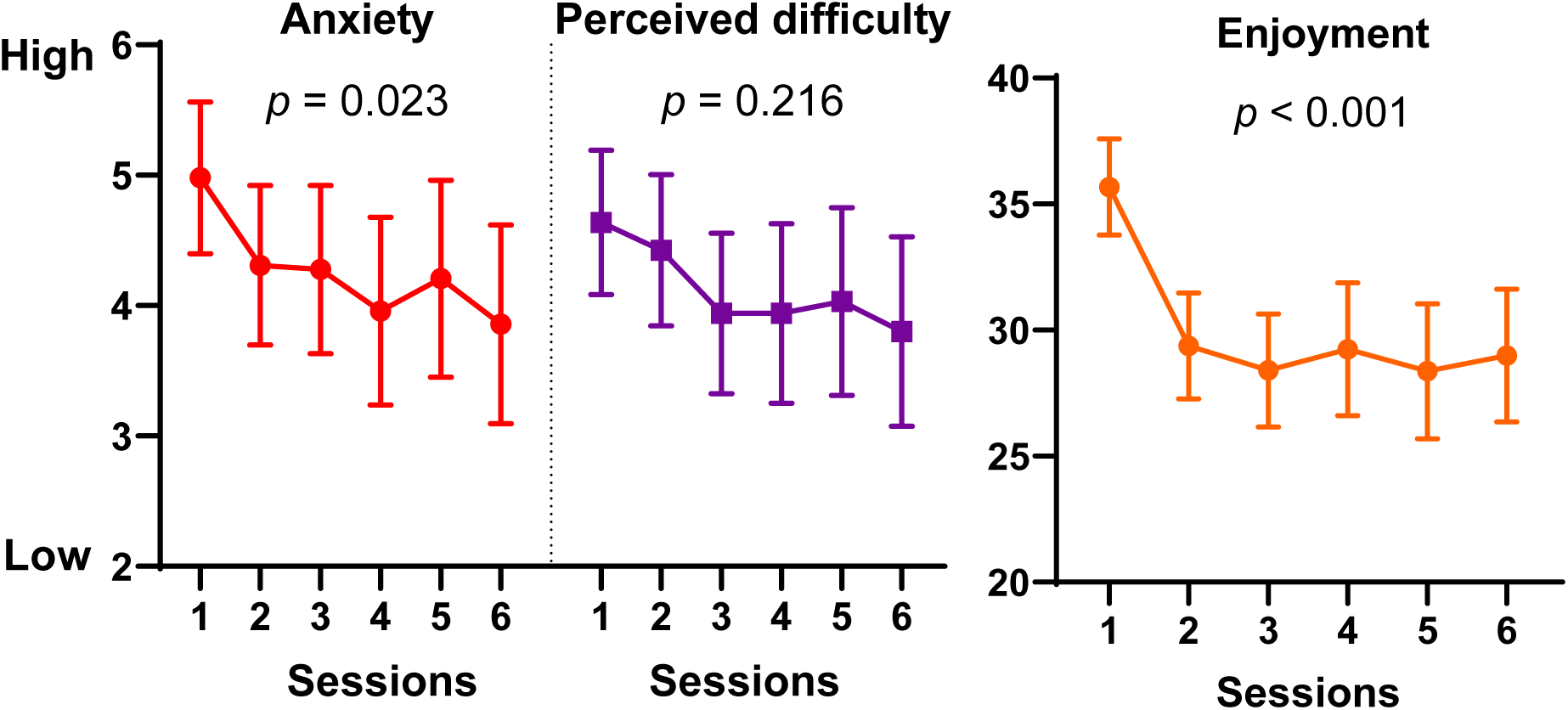
Subjective measures of participant’s anxiety, difficulty and enjoyment during the perturbation-based balance training using trips and slips. Sessions 1, 2, 3, 4, 5 and 6 correspond to weeks 1, 2, 3, months 3, 6 and 9, respectively. Linear mixed effects model was used to test if subjective measures changed over time. Fixed effects of sessions were tested using Type III FLtests. Points and error bars are estimated marginal means and their 95% confidence intervals.

### Laboratory induced trips and slips

At the 12-month reassessment, the intervention group had fewer laboratory slip falls than the control group (RR: 0.54, 95%CI: 0.30, 0.98, *P* = .043) with no significant difference found for laboratory trip falls (RR: 0.87, 95%CI: 0.64, 1.18, *P* = .367). Overall, the intervention group had fewer laboratory falls from the trip and slip (RR: 0.74, 95%CI: 0.54, 0.99, *P* = .040).

### Falls in daily life

In the ITT analyses (Figure 3), no significant between-group differences were observed in the overall number of trip and slip encounters, or in the number of trip- or slip-related falls (*P*>.07). The overall fall rate in daily life over one year was also not significantly different between the groups (RR: 0.77, 95% CI: 0.46, 1.30, *P* = .325). During the first three months, the intervention group experienced significantly fewer falls (RR: 0.38, 95% CI: 0.15, 0.97, *P* = .044), but this reduction was not maintained over subsequent follow-up periods (Q2–Q4) (*P* > .337). Significantly fewer injurious falls over one year were reported by the intervention compared to the control group (RR: 0.51, 95% CI: 0.29, 0.92, *P* = .024).

**Figure 3.**
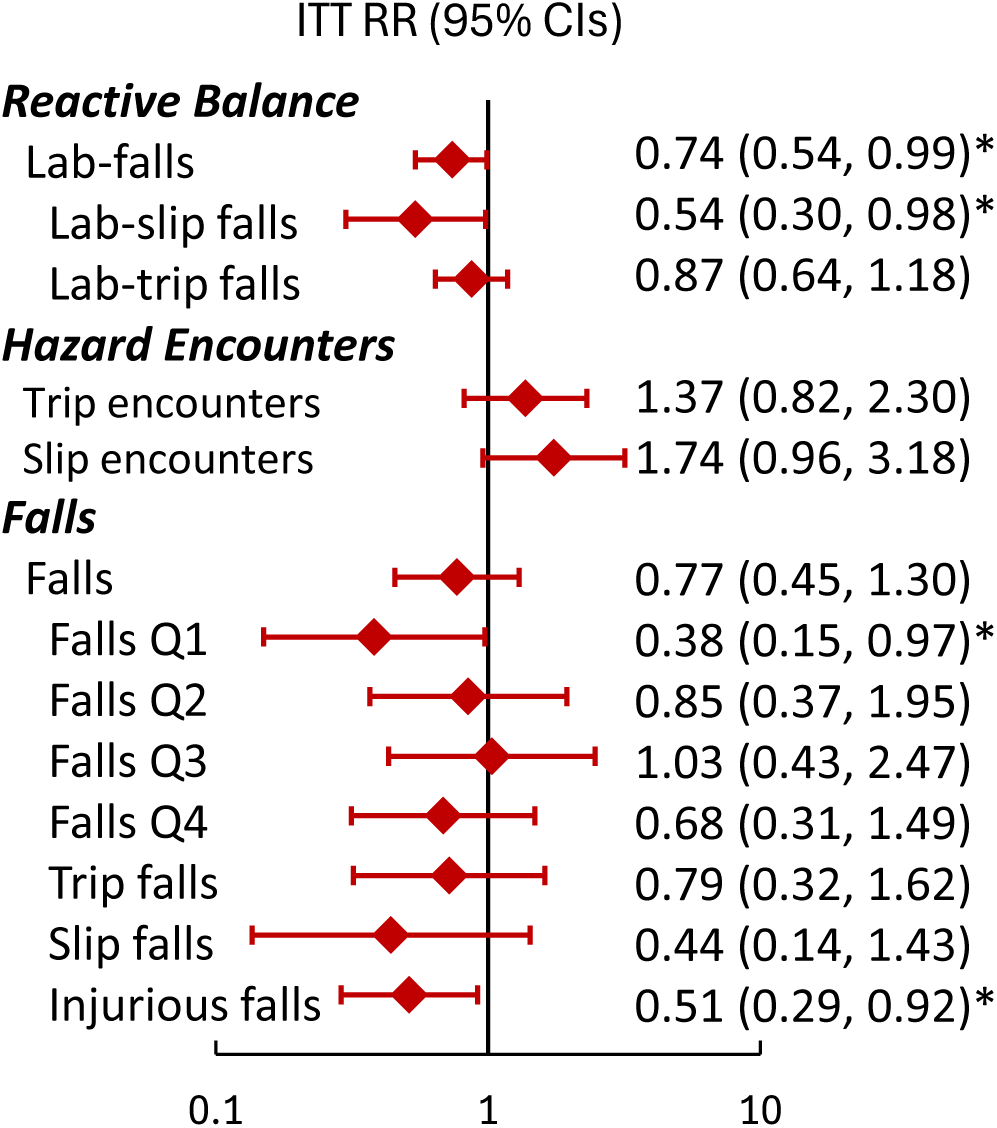
Rate ratios and 95% confidence intervals (95% CIs) of fall events in the intervention group relative to the control group, as intention-to-treat (ITT, n=111) analyses. To examine retention effects, one year was divided into quarters 1-4 (Q1-Q4). Trip and slip encounters include those resulted in a fall and recovery. * *P* < .05.

The CACE analysis showed a tendency toward greater reductions in fall rates during the first three months (RR_=_0.20, 95% CI_0.04–0.93) and in injurious falls over one year (RR_=_0.27, 95% CI_0.03–1.26), but not in overall falls over 12 months (RR_=_0.94, 95% CI_0.29–2.22).

### Functional, neuropsychological and behavioural factors

There were no significant differences between the groups at the 12-month reassessment for any of the functional, neuropsychological or behavioural factors (all *P* ≥ .062) (Table 2).

**Table 2.** Effects of the intervention on the secondary outcomes among the intervention group and control group.

| Variables | Baseline | | 12-month Reassessment | | *P* | $\eta^2$ |
| --- | --- | --- | --- | --- | --- | --- |
|  | Intervention | Control | Intervention | Control |  |  |
|  | (n = 54) | (n = 56) | (n = 29) | (n = 47) |  |  |
| Choice stepping reaction time (s) | 1.10 ± 0.21 | 1.04 ± 0.14 | 1.07 ± 0.16 | 1.05 ± 0.13 | 0.062 | 0.032 |
| Fall Behavioural Scale (FaB) (1-4) | 2.32 ± 0.33 | 2.31 ± 0.33 | 2.38 ± 0.33 | 2.34 ± 0.34 | 0.259 | 0.011 |
| Fall Efficacy Scale - International (FES-I)<br>(1-64) | 20.4 ± 4.26 | 20.4 ± 3.23 | 20.8 ± 3.55 | 19.8 ± 3.12 | 0.820 | 0.000 |
| Physiological fall risk (PPA) (z score) | 1.02 ± 0.86 | 0.81 ± 0.82 | 0.76 ± 0.82 | 0.69 ± 0.68 | 0.447 | 0.007 |
| General anxiety disorder (GAD7) (0-21) | 1.76 ± 2.99 | 1.71 ± 2.56 | 2.18 ± 3.04 | 2.00 ± 2.68 | 0.807 | 0.002 |
| Total physical activity (h/wk) | 32.8 ± 16.7 | 31.1 ± 17.5 | 35.1 ± 16.1 | 31.2 ± 19.5 | 0.526 | 0.006 |
| Trail Making Test B-A (s) | 47.7 ± 24.8 | 44.5 ± 21.8 | 51.5 ± 30.7 | 48.5 ± 22.0 | 0.778 | 0.000 |

$\eta^2$ of 0.01, 0.06 and 0.14 indicate small, medium and large effects, respectively. Higher PPA scores indicate higher fall risk. Higher GAD7 scores indicate greater anxiety. High FES-I scores indicate greater concern about falling. Higher FaB scores indicate safer behaviours. The table shows the observed number of participants but estimated marginal mean, standard deviation, *P* and $\eta^2$ are from models with multiple imputation.

## DISCUSSION

This assessor- and analyst-blinded RCT found a 12-month PBT program improved reactive balance and reduced injurious falls in community-dwelling older adults. Notably, this effect was achieved with a relatively low overall training dose, consisting of six sessions in total: three initial weekly PBT sessions followed by three re-training sessions delivered at three-month intervals. Improvements in reactive balance were evident following slip perturbations, which were reduced by 46% in the intervention group compared to the control group. Furthermore, total falls during the first three months decreased by 62%, demonstrating substantial short-term benefits of PBT. These findings partially support our hypothesis and offer valuable insights for integrating PBT into fall prevention programs.

This RCT extends the evidence that short-term slip- and trip-based PBT enhances reactive balance, ^23^ ^39^ and demonstrates its effects on balance recovery in the medium- and long-term. Consistent with previous findings,^23^ ^40^ PBT improved reactive balance over 12 months and reduced fall rates during the first 3 months among participants who completed the initial three weekly sessions (Figure 3). However, the repeated practice through three-monthly re-training sessions, did not maintain the reduction in fall rates over the remaining nine months, with no differences observed between the intervention and control groups at the 12-month reassessment. This is consistent with prior studies suggesting that adaptations to PBT may diminish within three months without sufficient booster sessions to reinforce previously acquired sensorimotor skills^15^ ^24^, but contrasts with the findings of Bhatt 2009 who reported that high-intensity PBT, involving many repeated perturbations, may be sufficient to sustain improvements in reactive balance (e.g., laboratory falls).^41^ Attrition is also likely to have influenced the efficacy of the intervention, as most withdrawals occurred within the first three months, and COVID-19 lockdowns in 2020 and 2021 greatly disrupted the scheduling of the re-training sessions. Overall, these findings highlight the importance of short-term PBT in establishing a protective effect against falls and suggest that greater training stimulus, through higher intensity (e.g., unpredictability) or more frequent re-training sessions, may be required to sustain benefits over time.

Contrary to our hypothesis, no differences in functional, neuropsychological, behavioural or cognitive fall risk factors were observed between the intervention and control groups at the end of the trial. Similar findings have been reported following a treadmill PBT program,^42^ and attributed to the recruitment of healthy participants. Taken together, these findings suggest that PBT is a highly task-specific intervention which effectively targets reactive balance in a manner that can be generalised to real-world scenarios. It is likely that the mechanisms underlying the improved reactive balance and reduced fall injuries from PBT lie in the improved efficiency of coordinating muscle and kinematic responses.^43 44^ Furthermore, the significant reduction in injurious falls may indicate that PBT enables individuals to implement protective strategies when a fall does occur, thereby reducing the likelihood of fall-related injuries. Our findings also suggest slip perturbations are more responsive to a mixed model of trip- and slip-based PBT. This may reflect consistency in practice, as slip exposure consistently occurred at foot-strike, whereas trip recovery is more complex, requiring strategy selection (elevating or lowering) depending on the gait phase (early, mid, or late swing) at obstacle contact.^45 46^ Although no significant effect was observed on overall fall rates, the re-training sessions appeared sufficient to preserve some benefits in reactive balance, as evidenced by a 57% reduction in injurious falls and a 26% reduction in laboratory falls at the 12-month reassessment in the intervention group compared with controls.

The efficacy of PBT, characterised by a low dose yielding a large response, is considered a key advantage over conventional exercise programs. In this RCT, initial adherence to the PBT program was high (85%), but dropout from booster sessions was substantial (46%), likely influenced by psychological effects following perturbation exposure, including anxiety (Figure 2). Although anxiety decreased with repeated sessions and participants were reassured that physical risks were minimized through padding and harness use, the simulation of real-life fall circumstances may have contributed to apprehension during training. Protocol adjustments, shifting from unpredictable perturbations at program onset to a graded approach beginning with predictable training, resulted in only marginal improvements in dropout rates (47% pre-adjustment vs. 34% post-adjustment).

These findings highlight several directions for future research. First, the acceptability of a progressive, individualised approach to better support participants and enhance reactive balance should be evaluated. Second, further work is needed to develop and validate PBT programs for clinical settings. While commercially available perturbation treadmills or manual techniques may be feasible, evidence suggests that treadmills do not sufficiently destabilise or train the stepping responses required to clear trip obstacles encountered in daily life^17^ ^18^ ^19^ Third, optimal PBT parameters (e.g. training intensity and dosage) remain unclear, given the multiple avenues for progression such as gait speed, obstacle height, timing within the gait cycle, and slip distance. Finally, populations most likely to benefit from PBT must be identified. Our findings suggest that PBT may be particularly suitable for healthy adults. Addressing these questions will be critical for the large-scale implementation of PBT in fall prevention.

### Clinical implications

Our findings support the integration of task-specific PBT into fall prevention programs for older adults, as conventional strength and balance programs do not sufficiently address reactive balance control. A short, structured PBT program could complement existing interventions and provide a time-efficient strategy to reduce serious fall risk in older populations. Importantly, such programs should adopt a tailored approach to appropriately challenge balance and incorporate frequent re-training sessions to ensure the retention of adaptations.

### Strengths and limitations of the study

This study had several strengths, including the use of blinded assessors, rigorous fall data collection through weekly SMS surveys with a 95% response rate, and objective verification of outcomes. However, certain limitations should be acknowledged. The relatively healthy and motivated sample may limit generalisability to frail or sedentary populations. Moreover, the study was conducted during the COVID-19 pandemic, which led to recruitment challenges, disruptions to the re-training schedule, and a considerable number of dropouts. These factors likely contributed to the study being underpowered and may have influenced the long-term outcomes.

### Conclusions

A brief PBT program involving repeated trips and slips, supported by periodic booster sessions improved reactive balance at 12 months and reduced injurious falls by 57% in older adults. Trip- and slip-encounters did not change, indicating that benefits were likely due to enhanced reactive balance rather than proactive gait strategies. Further research is needed to establish optimal dosing, delivery methods, and implementation strategies to enhance real-world impact, long-term retention and scalability.

## ACKNOWLEDGEMENTS

We acknowledge Ms Natassia Smith, Courtney Lee, Cathy Jung, Maoling Lim, Hannah Gibson, Carmen Tung and Mr Key Nahan for their contribution as research assistants, blinded assessors and trainers for this SafeTrip trial. We sincerely thank A/Prof Roberta Shephard for advising us as a consumer representative, Mr Hilary Carter and Mr Artemij Iberzanov for their contribution as mechanical and electrical engineers. We express our deep gratitude to all participants who volunteered to take part in this SafeTrip trial.

## Author Contributions

Y.O., S.P., C.C. and S.R.L. contributed to the conception and design of the study. S.P., C.C., Y.O., C.H., E.C. and S.P. administrated the study. C.H. and P.H. conducted the statistical analyses. The manuscript was drafted by Y.O. and reviewed and edited by S.P., C.C., C.H., E.C., P.H. and S.R.L.

## Data availability statement

Requests for de_identified participant data, including assessment results collected during the trial, should be directed to the Principal Investigator and the UNSW Human Research Ethics Committee. Access to data will be considered on a case_by_case basis in accordance with ethical and institutional requirements.

## Patient and public involvement

An older adult with a physiotherapy background contributed to this trial as a consumer representative, participating in and providing feedback during the refinement of the reactive balance training program.

